# Field study to determine the reliability of HIV viral load results shows minimal impact of delayed testing

**DOI:** 10.1101/2023.08.30.23294838

**Authors:** Diana Hardie, Howard Newman, Joanna Ried, Nei-Yuan Hsiao, Gert van Zyl, Lucia Hans, Jasantha Odayar, Stephen Korsman

## Abstract

Understanding factors that impact on the accuracy of HIV viral load (VL) results is key to quality improvements of VL programmes, particularly in resource limited settings. In this study we evaluated whether testing delay and room temperature storage (between 25-30°C) prior to testing affected results. 249 individuals who were on antiretroviral therapy, or with newly diagnosed HIV, were recruited and three PPT samples were collected from each patient. One sample was tested within 24 hours, while the others were stored un-centrifuged at ambient temperature prior to testing.

Centrifugation and testing of matched samples were performed on days 4 and 7 after collection. In samples with initial VL >2 log copies/mL, time delay and ambient temperature storage had minimal impact. When grouped according to day 1 VL range, 96-100% of samples at day 4 and 93-100% at day 7 had a VL within 0.5 log copies/mL of the first result. Greatest variability was observed at day 4 and 7 when the day 1 VL was <2 log copies/mL, however, there was no trend of increasing difference with time. Of day 1 samples with undetectable VL or VL <50copies/mL, 80% and 78% had concordant results at days 4 and 7, respectively. Detection in later samples appeared to be stochastic, rather than showing a worsening trend. These results provide further evidence that VL is stable in PPT tubes for up to 7 days when stored at room temperature. It shows that there is significant variability in samples with VL close to the limit of detection, not affected by testing delay. Stochastic variation or technical factors that may increase the chance of false positive results could possibly account for this.

## Introduction

Viral load (VL) testing is a key element to reaching and monitoring progress towards the UNAIDS 95:95:95 targets(1). This reflects ensuring that HIV-infected patients are diagnosed, started on anti-retroviral therapy (ART) and are virologically suppressed. South Africa (SA) has the world’s largest treatment program with > 6 million people living with HIV (PLWH) enrolled(2). Over 6 million viral loads were performed by public sector laboratories in SA in 2022(3). However, logistics and pre-analytical challenges such as transporting, registering, centrifuging and storing samples could compromise the quality of testing.

Currently manufacturers of high throughput VL platforms such as Roche Molecular Systems (Pleasanton, USA) and Abbott Laboratories (Chicago, USA) recommend that whole blood samples should be centrifuged within 6 to 24 hours of collection and plasma should either be frozen at - 80 ^⁰^C or tested within 5 days of sample collection(4)(5). Transporting samples to the laboratory for separation into plasma is often not achievable within the official manufacturer time constraints in resource-limited settings. Samples may reach the laboratory more than 24 hours after collection and are stored at 4°C in their primary tubes until testing. The extent to which these pre-analytical factors compromise the accuracy of results was assessed in two laboratory simulation studies (6)(7) and a systematic review (8). All three showed good preservation of viral RNA beyond the currently recommended testing time. This finding was confirmed in a local study performed at Groote Schuur Hospital in which routine diagnostic samples collected in ethylenediaminetetraacetic acid (EDTA) tubes or EDTA-plasma preparation tubes (PPT) were stored after initial HIV VL testing at a range of times and storage temperatures(9). The VL in samples stored for up to a week reliably differentiated between ART-suppressed and failing patients in 98.8% of instances(9). However, samples had already been centrifuged prior to storage and the study was performed entirely in the laboratory environment. In addition, it was only possible to follow trends in VL from the same patient over 2 time points due to sample volume constraints. In this current study, we evaluated the extent to which testing delay and adverse sample storage affects VL by testing serial samples from the same patients at different time points after sample collection.

### Study objectives

1. To determine the impact of testing delay on the accuracy of HIV RNA quantification in diagnostic specimens collected and stored in un-centrifuged PPT tubes.
2. To describe the impact of storage at a warmer ambient temperature (storage between 25-30°C) on the precision of HIV RNA quantification in diagnostic samples in un-centrifuged PPT tubes.
3. To determine whether centrifugation prior to storage improves the precision of results in samples where testing is delayed and whether omitting a 2^nd^ re-centrifugation step prior to delayed testing affects precision (evaluated at one site, namely GSH).

## Methods

Ethical approval for the study was obtained from the Human Subjects Research Ethics Committee of the Faculty of Health sciences at the University of Cape Town (HREC Ref 159/2019). Written informed consent was obtained from all participants prior to enrolment. Only individuals older than 18 years were recruited.

Enrolment for 2 sites (PE and TBH) began on 21 November and 5 December 2019 and ended on 24 January and 23 January 2020, respectively. Enrolment for the 3^rd^ site (GSH) began on 28 August 2022 and ended on 6 June 2023 (following normalisation of laboratory services after SARS-CoV-2 pandemic).

The study was performed at three sites within the National Health Laboratory Service (NHLS) network of HIV VL testing laboratories. Patients were recruited from four ARV clinics, three in the Cape Metro and one in the Eastern Cape Province of South Africa. Patients who were due to have a VL test or who were newly diagnosed and not yet on therapy, were approached and asked to give two extra blood tubes for HIV VL testing. Three PPT samples were collected during the same blood draw and delivered to the regional HIV VL testing laboratory. On arrival, one sample was centrifuged and sent for immediate HIV VL testing. Testing was completed within 24 hours of sample collection. The result from this test was issued for routine patient management. The other two tubes were stored un-centrifuged in routine field conditions where the temperature varied between 25-30°C. Four days after sample collection, one of the stored samples was centrifuged and a VL test was performed. Seven days after sample collection, the third tube was centrifuged and tested. VL testing was performed using the routine assay in each of the three participating laboratories at the time of study, namely Roche CAP/CTM (Tygerberg laboratory), Roche 6800 (Port Elizabeth laboratory) and Abbott Alinity m (Groote Schuur laboratory). The study plan is given in Fig 1.

**Fig 1:**
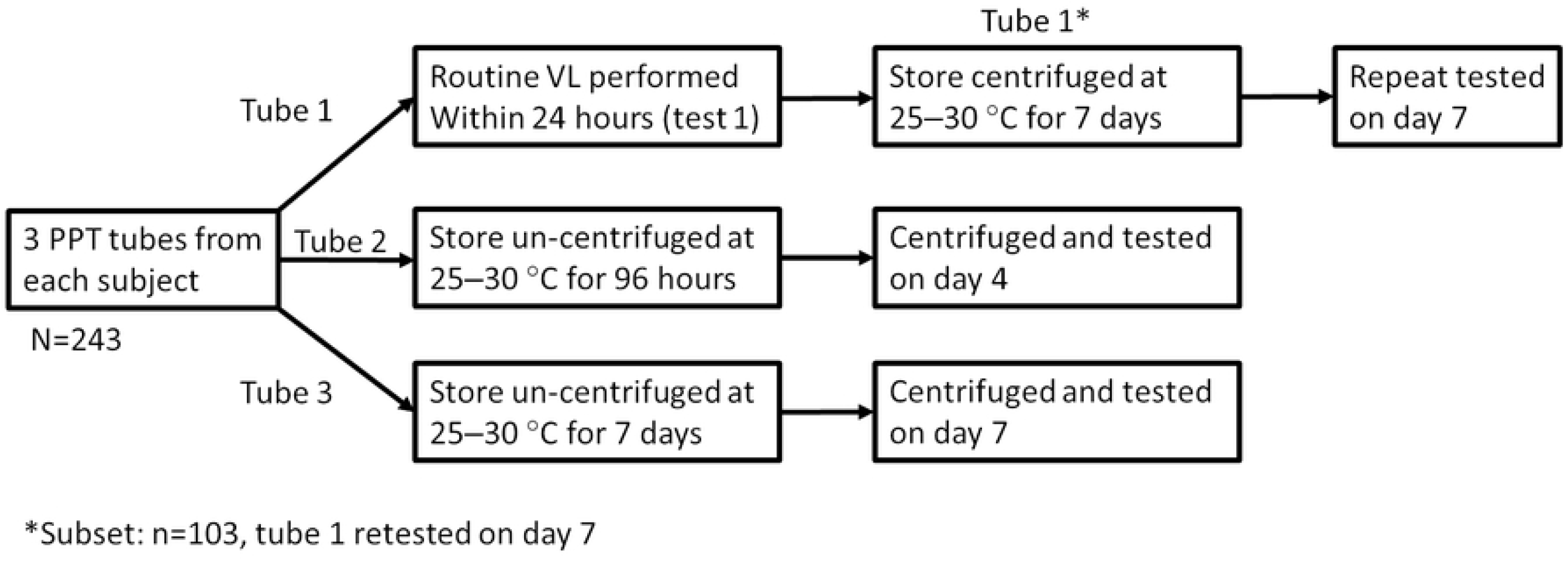
Study plan

On a subset of patients (done at Groote Schuur laboratory only), HIV VL was repeated at day 7 on the sample that had been centrifuged and tested on day one. The day 1 centrifuged samples were stored under the same conditions as the yet untested samples, prior to repeat testing. The samples were not re-centrifuged prior to repeat testing on day 7. They were used to assess whether storage in an already centrifuged state improved stability of VL in the sample.

VL data from days 1, 4 and 7 testing were extracted from the laboratory information system and analysed using Bland-Altman plots (10).

For analysis, unless otherwise specified, samples with a VL that was lower than the limit of detection (LDL) were assigned a value of”1” (0 log) copies/mL. Samples with a VL value of <20 copies/mL, were assigned a value of”19” (1.28 log) copies/mL, otherwise the log value of the reading in copies/mL was used for analysis.

A difference in VL of >0.5 log copies/mL was considered significant and likely to reflect a change that was caused by factors other than random expected variation. (11)

## Results

A total of 249 patients were consented and provided 3 PPT samples for testing (51 from Tygerberg, 60 from Port Elizabeth (PE) and 138 from Groote Schuur (GSH). Samples from 6 patients arrived at the laboratory more than 24 hours after collection and these patients were excluded from the study.

Day 1 VLs segregated into the following VL categories: 55 (22.6%) were LDL, 46 (18.9%) were detectable, but <50 copies/mL, 7 (2.9%) were 50-100 copies/mL, 28 (11.5%) were 2-3 log copies/mL, 30 (12.3%) were 3-4log copies/mL, 36 (14.8%) were 4-5log copies/mL, 30 (12.3%) were 5-6log copies/mL and 11 (4.5%) were >6 log copies/mL.

### Viral load stability at day 4 and 7

Bland-Altman plots were used to evaluate differences in day 1 vs day 4 VL results and between day 1 and day 7(10). Overall, there was very little difference in paired readings at both day 4 and day 7 compared with day 1. The mean bias at day 4 was -0.15 and for day 7 was -0.12 log copies per mL, indicating a lower reading in the samples where testing was delayed, but the decay was minimal **(Fig 2**). For samples with an initial VL of >2 log copies/mL, the paired VL value at day 4 and day 7 was within 0.5 log copies/mL in 96-100% of instances for day 4 and 93-100% of instances for day 7 (**table 1**), signifying minimal impact of the testing delay on the quantification. The greatest variation was observed for samples with day 1 VLs less than 2 log copies/mL, with only 60% of samples at day 4 and 62% at day 7 having a repeat viral load within 0.5 log copies/mL. In this very low viraemic range, even though many of the changes were >0.5 log copies/mL, the difference in quantification does not reflect real clinically significant differences in VL value. No clinical action would be indicated for results in this (<2 log copies/mL) range(12)(13). Indeed, in the 101 samples with a day 1 viral load below 50 copies/mL, 80% remained <50 copies/ml at the day 4 test and 78% at the day 7 test.

**Table 1:** Proportion of samples tested on day 4 and 7 within 0.3 and 0.5 log copies/mL of day 1 viral load value by day 1 VL category:

| Day 1 viral load by category | Number of samples | Day 4 viral load % within 0.3 log of day 1 result | Day 4 viral load % within 0.5 log of day 1 result | Day 7 viral load % within 0.3 log of day 1 result | Day 7 viral load % within 0.5 log of day 1 result |
| --- | --- | --- | --- | --- | --- |
| LDL | 55 | 51% | 51% | 60% | 60% |
| <log2 | 53 | 51% | 60% | 55% | 62% |
| Log2-3 | 27 | 89% | 96% | 81% | 93% |
| Log3-4 | 30 | 87% | 97% | 87% | 97% |
| Log4-5 | 36 | 81% | 100% | 86% | 97% |
| Log5-6 | 30 | 93% | 97% | 97% | 97% |
| >log6 | 11 | 73% | 100% | 91% | 100% |

**Figure 2:**
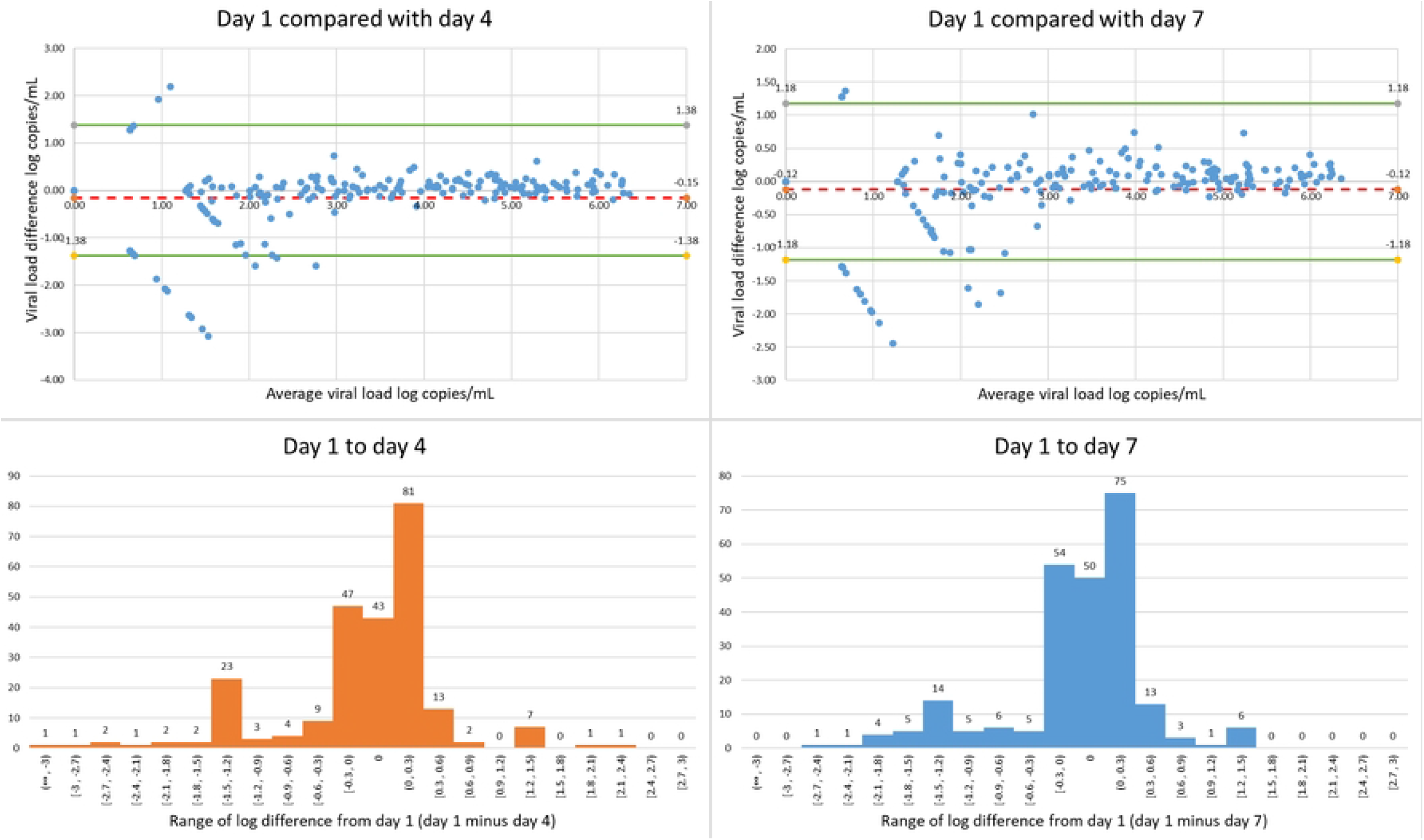
Variability of all paired VL results at day 4 and 7: Bland-Altman plots of day1 vs day4 VL (a) and day1 vs day7 (b) show negative bias of -0,14 log copies/mL at day4 and -0,12 log copies/mL at day 7, with greatest variability at lower day 1 VL (<2logcopies/mL). Green lines represent + and -2SD and red dotted line the mean difference in HIV viral load on Bland Altman plots. Histograms of paired VL differences at day 4 and 7 are shown in(c) and (d). The peaks at -1.2 to -0.9 and +0.9 to 1.2 mostly reflect samples with VLs that were <1.3 log copies/mL which transitioned to LDL or LDL samples that transitioned to <1.3log copies/mL.

### Viral load precision between 100-1000 copies/mL (2-3 log copies/mL)

27 samples had day 1 VL values between 100 and 1000 copies per mL. Variability was much less, with 96% of samples on day 4 and 93% on day 7 having VL within 0.5 log copies/mL. The box and whisker plot **(Fig 3a)** which included all samples with quantifiable VL from 20-1000 copies/mL show the overlapping VL ranges at day 1, 4 and 7, with occasional outlier values.

**Figure 3:**
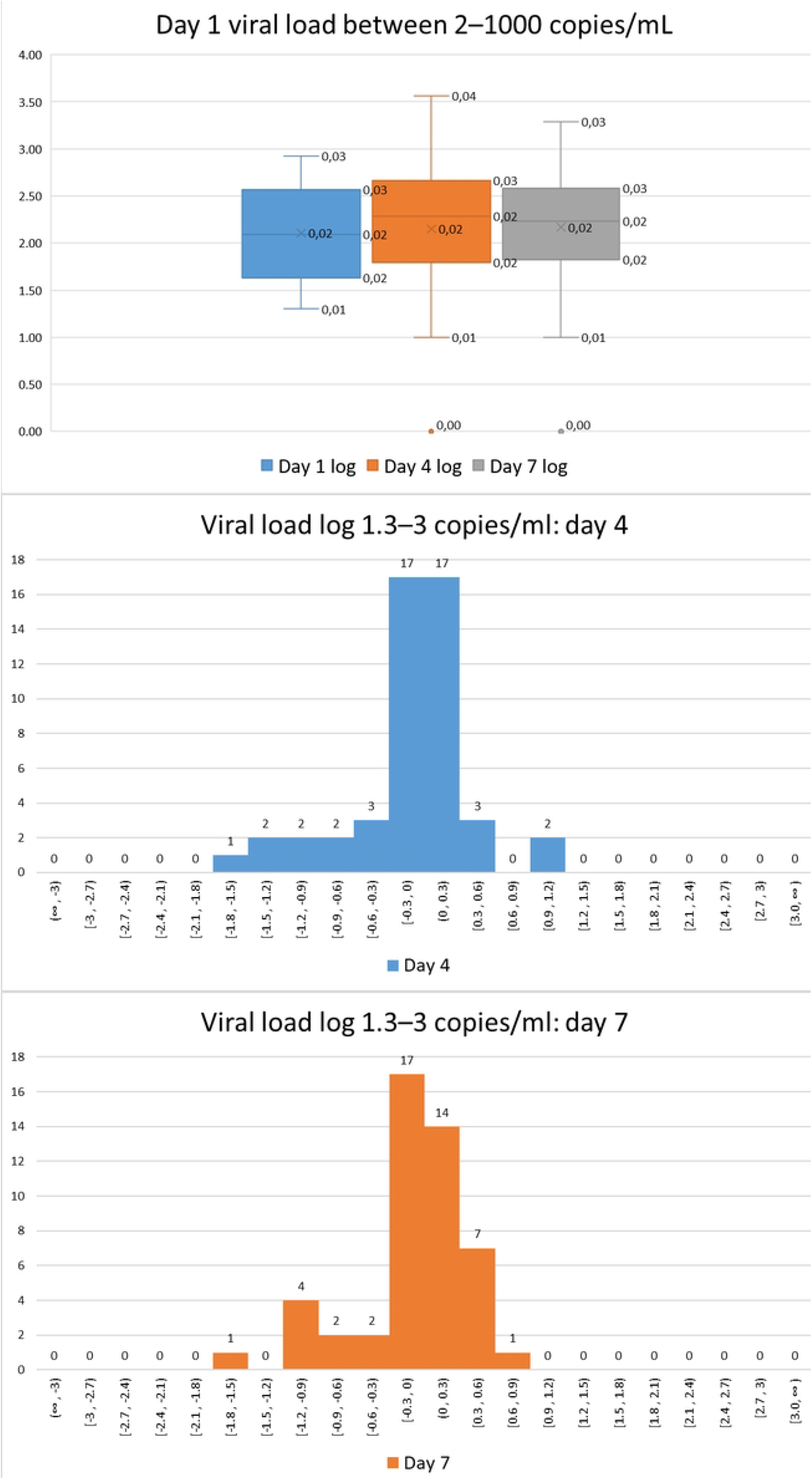
Viral load changes in samples with day 1 viral load between 1.3-3 log copies/mL: (a) shows a box and whisker plot comparing viral load ranges at 1, 4 and 7 days. Histograms in (b) and (c) show the variation in paired samples at day 4 and 7.

### Day 1 undetectable/<50 copies/mL samples that became detectable on repeat testing

In total, 18 patients who had viral loads <50 copies/mL at day 1 had a repeat viral load of >2 log copies/mL at day 4 or day 7 **(Fig 4a)** and of these, 8 were >2 log copies/mL at both time points. Detection in a later sample appeared to be stochastic. There was no trend to suggest that this phenomenon worsened with time. The testing site or automated technology used to perform testing seemed to be an important factor as this varied from 0% (GSH, Abbott Alinity), 11% and 22% at days 4 and 7 (Tygerberg, Roche CAP/CTM) to 50% (PE, Roche Cobas 6800). (**Fig 4b**)

Patients with a day 1 VL of detectable, but <20, were 2.3 times more likely to have a detectable VL at day 4 and 1.8 times more likely to have a detectable VL at day 7 than patients with an undetectable day 1 VL (LDL).

**Figure 4.**
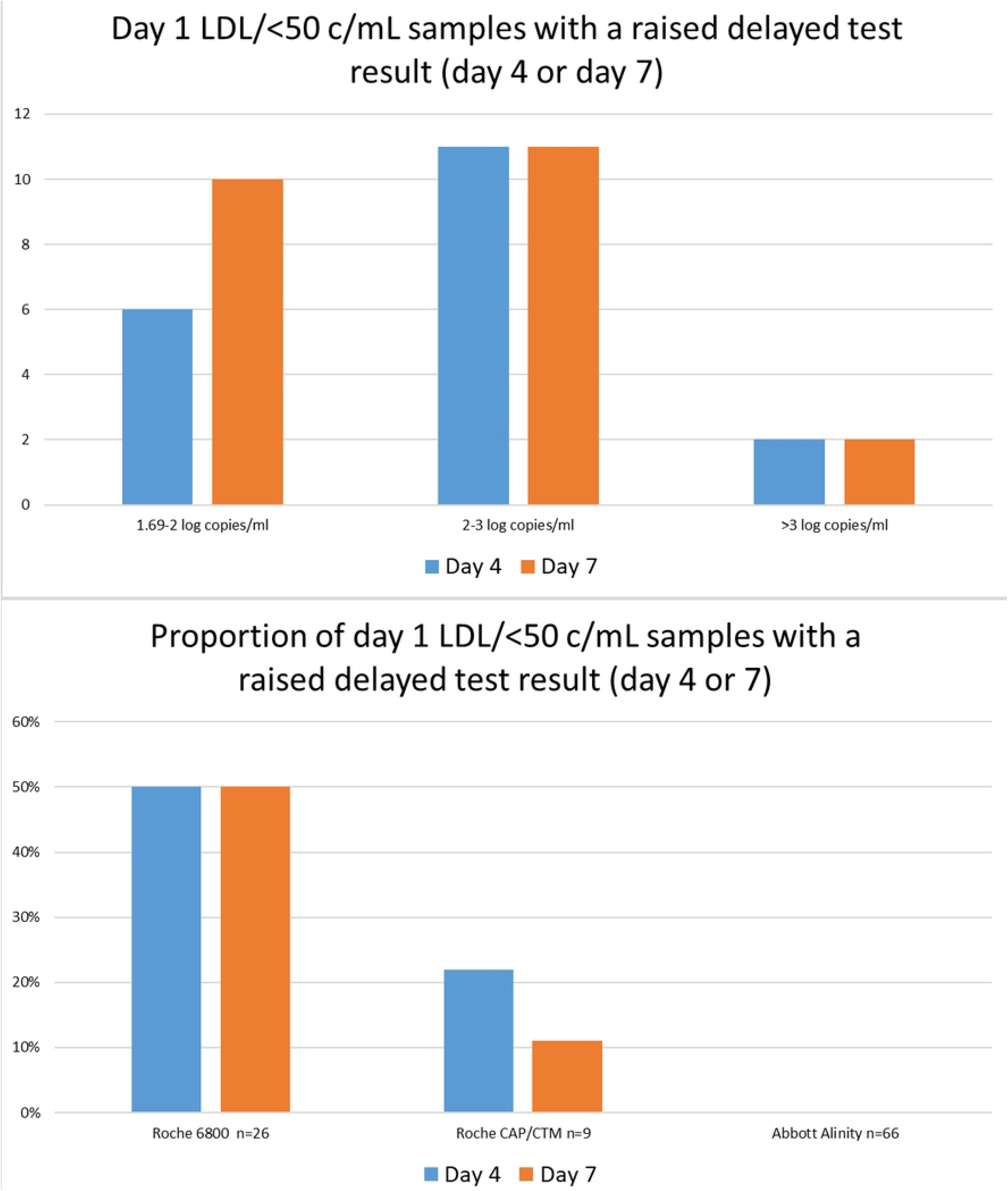
(a) shows the number and viral load ranges on days 4 and 7 of samples that were day 1 LDL or had values <50 copies/mL which became detectable on repeat testing and (b) shows the proportion of samples with day 1 viral loads of LDL or <50 copies/mL that had a VL >2 log copies/ml on repeat testing, by testing site/technology used.

### Stability in centrifuged samples

VL stability in samples that were centrifuged on day 1 and (re-) tested on day 7 was also analysed in a subset of 103 samples from one centre (GSH). The day 1 sample was stored after initial testing under the same conditions as the un-centrifuged samples and was retested on day 7 (without recentrifugation). The VL values on day 1 and 7 were very similar (**Fig 5**). The mean difference in VL between day 1 and day 7 was 0.03 log copies/mL and SD was narrower at 0.42 than for the samples that were first centrifuged and tested on day 7. No samples had a clinically significant change in VL when comparing the day 1 and day 7 samples. One sample had an initial VL of 4.95 log copies/mL which changed to 3.85 log copies/mL. Thus, there was only one result in this group with a clinically significant (-1.1 log copies/mL) VL change. The good results in this experiment could reflect: better sample preservation during storage when sample is separated on day 1, lack of viral RNA leakage in the PPT tubes on storage (previously flagged as a potential problem (14)(15)) or other technical factors relating to the testing site.

**Figure 5:**
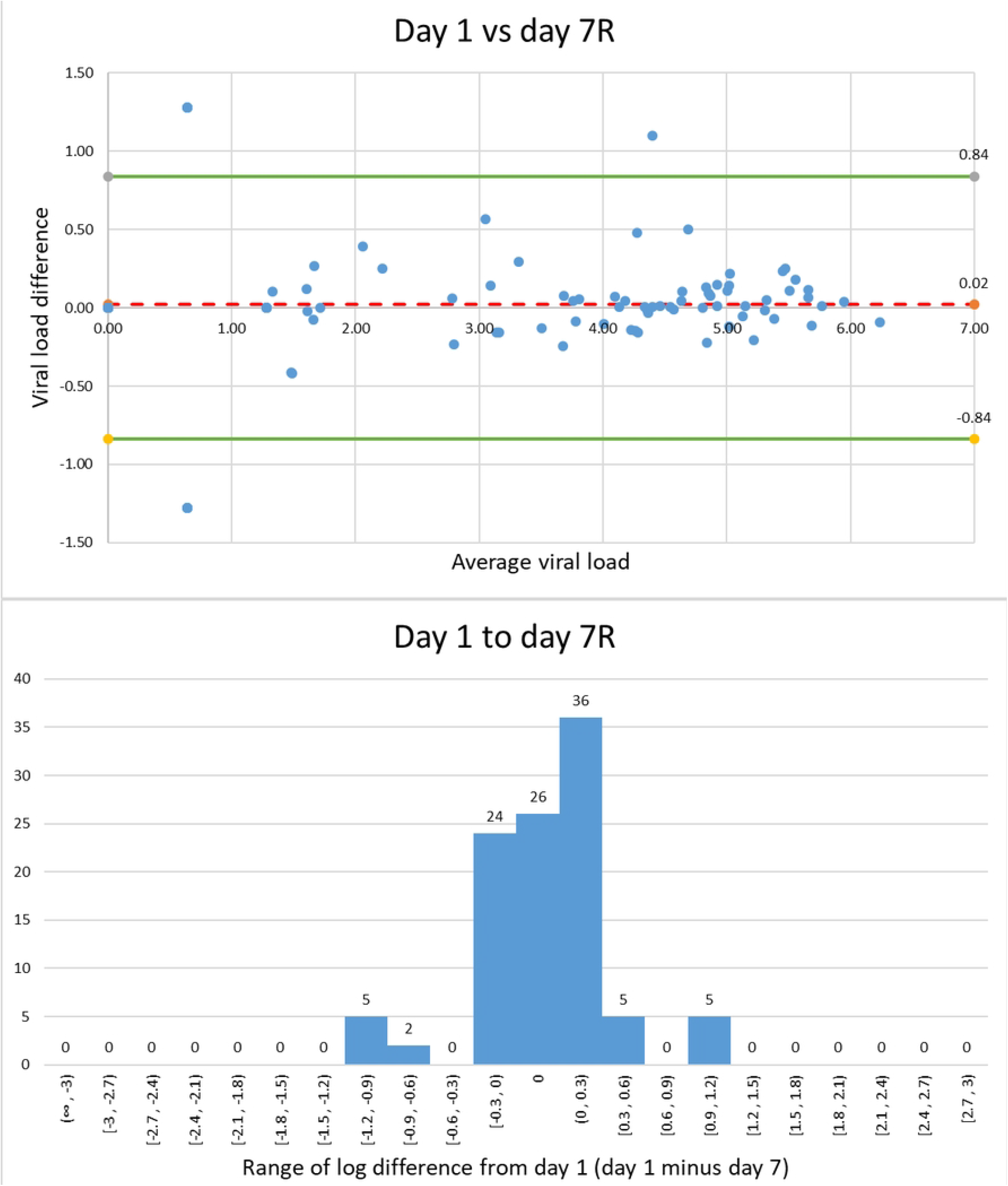
Variability of paired viral loads of day 1 samples that were re-run on day 7: (a) Bland Altman plot reveals minimal bias and lower variance in day 1 vs day 7 results. Green lines represent + and - 2SD and the red dotted line the mean difference in HIV viral load. Histogram in (b) shows the difference in day 1 and 7 viral load results.

## Discussion

Various factors impact on the accuracy of HIV VL test results (lucia’s paper) and an understanding of these is key to better interpretation of the result and mitigation of potential inaccuracies. In this study we found that delay of testing for the first 7 days had minimal effect on test results. This was clearly shown for samples with an initial VL >2 logs. Testing of paired samples at day 4 and 7 returned equivalent results in 96-100% (at day 4) and 93-100% (at day 7) of instances when stratified by VL level at day 1 (sample tested within 24 hours of collection). These findings are compatible with previous work done by ourselves (9) and a systematic review by Bonner et al (8) which showed that VL was stable when samples are stored under laboratory conditions for up to 7 days. However, here we provide additional field data on the stability of samples with a day 1 VL below 3 log copies/mL and also show that storage of samples in an un-centrifuged state are stable when stored at ambient temperatures between 25 and 30°C for up to 7 days. This more closely matches the real-world situation, especially in low-and-middle-income countries with high HIV disease burdens and large antiretroviral treatment programmes such as in SA. Under these conditions testing is often delayed due to logistical issues, especially in remote rural areas, and these results provide reassurance that time delay is not a major cause of result inaccuracies.

Significant variability was observed in samples with day 1 VLs of <2log, with only 60% (on day 4) and 62% (on day 7) of samples having repeat VL values within 0.5 log copies/mL. But factors other than time delay are more likely responsible. Variability in logarithmic terms near the limit of detection is expected to be greater because at this level, detection/loss of detection and outlier results will represent a bigger logarithmic change(16). Clinicians should be aware of the inherent VL variability in this range and interpret results with caution.

Both local and international guidelines advise using VL levels of <50 copies/mL to reflect viral suppression in patients on antiretroviral therapy (ART) (12)(17) and it was reassuring that for samples with a day 1 VL in this range, 80% at day 4 and 78% at day 7 remained in this category on repeat testing. Nonetheless, recent studies have highlighted the diagnostic importance of persistent low level viraemia in patients on ART as a marker of increased risk of virological failure(18,19) and a better understanding of the cause of these low positive results is needed.

Low VLs could also result from detection of intra-cellular viral nucleic acids in stored samples (12,13). Sometimes cellular material remains above the gel in PPT tubes after centrifugation (15)and aspiration during the testing process can falsely elevate the VL in recently suppressed individuals(20). This phenomenon is only likely to make a difference if the plasma VL is very low or undetectable to start off with. This could account for some of the 20% of patients who had suppressed (<50 copies/mL) day 1 VLs, that were detectable in day 4 or 7 samples. Of note, samples from patients that were detectable, but VL was <20 copies/mL at day 1, were twice as likely to have a detectable VL in a stored sample than patients that had an undetectable VL at day 1.

Noteworthy was the observation that the proportion of samples affected, varied markedly at the different testing sites. Technical factors such as duration and speed of centrifugation of primary samples, the method of sample aspiration by the auto-analyser or other undefined laboratory factors could increase false reactivity during the testing process. Re-centrifugation immediately prior to testing has been recommended (15) as a means to reduce false reactivity in stored samples, the rationale is that cellular elements containing viral nucleic acids may diffuse into the plasma when storage is prolonged.

In the subset of 103 patients evaluated, re-centrifugation was omitted when the sample first tested on day 1 was rerun on day 7. Interestingly, no samples had a clinically significant change in VL when comparing the day 1 and day 7 samples. This result was reassuring. A caveat is that this experiment was only done at the site where there was very low variability in the stored samples anyway.

## Limitations

Only three samples were collected from each patient, and this limited the number of factors we could evaluate. Testing occurred across 3 sites and different testing platforms could have accounted for some of the differences that were seen, for example, a much lower rate of detectable viral load in later samples with day 1 suppressed VL at one site. In the LMIC context where samples may arrive for testing after transport at temperatures above 30⁰C and in EDTA tubes without gel separator, more evaluation is needed to define the limits of acceptable pre-analytical conditions.

## Conclusion

Our field study provides further evidence that time delay had minimal impact on viral load quantification when samples were stored at room temperature. When the D1, D4 and D7 results were compared, substantial variability was observed in samples with VLs close to the limit of detection, but there was no trend to suggest increasing variance with time. Some samples with negligible VL at day 1 had detectable VL on samples tested later and this was more likely to happen when the day 1 plasma VL was <20 rather than LDL. The most likely explanation is that this reflects a combination of stochastic variation and false detection of low level viraemia during the testing process. The variation observed near the limit of assay detection could also contribute to the phenomenon of viral blips. The contribution of other patient-related and technical issues requires further investigation.

## Data Availability

file containing raw HIV viral load data has been included in supplementary data

## Supporting information

**S1 File_XLS: Raw data file containing day 1, 4 and 7 viral load results of study samples**.

